# Agentic Pipeline for Large-Scale Clinical Data Queries in Enterprise Electronic Health Records

**DOI:** 10.64898/2026.11.05.26367378

**Authors:** Kshitij Pawar, Nazgol Tavabi, Anvita Kalepalli, Nishita Kalepalli, Mallory Blackmon, Ashton Williamson, Ata M. Kiapour

## Abstract

Using electronic health record (EHR) data to answer clinical research questions is challenging. It requires understanding the research question, finding relevant information across different sources, and combining that information to produce meaningful results. We developed an agentic large language model (LLM) pipeline that decomposes investigator-authored requests into source-specific tasks, generates and executes SQL on EHR, integrates structured and unstructured EHR evidence, and applies study-specific clinical note adjudication. We evaluated this workflow against data analyst–identified cohorts across 50 pediatric orthopedic clinical data query requests. Of these, 43 were included in manual-review-adjusted performance analyses. The median adjusted positive predictive value, sensitivity among patients identified by either approach, and corresponding F1 score were 0.93 (95% CI, 0.88–0.98), 0.92 (95% CI, 0.90–0.95) and 0.90 (95% CI, 0.87–0.94), respectively. These findings support the feasibility of translating natural-language research intent into executable, autonomous multi-stage EHR workflows.

---

Clinical research requests often describe complex eligibility criteria in natural language that cannot be queried directly against electronic health record (EHR) data [1]. Staff must translate each request into computable rules, choose the relevant data sources, write institution-specific queries, and interpret the results. This work takes time and specialized expertise, yet supports feasibility assessment, retrospective studies, quality improvement, cohort instruction, and clinical trial recruitment.

Enterprise EHRs store evidence in both structured fields and free text. Structured data include diagnosis and procedure codes, laboratory results, medications, and imaging metadata. Clinical notes and imaging interpretations are unstructured. Important study criteria—such as diagnostic confirmation, severity, device use, laterality, and study-specific exclusions—may appear only in free text. Figure 1 shows this challenge. The request seeks pediatric patients with scoliosis treated using distraction-based growth-friendly surgery: magnetically controlled growing rods (MCGR), traditional growing rods (TGR), or vertical expandable prosthetic titanium ribs (VEPTR). Treatment, rather than scoliosis or spinal instrumentation alone, defines eligibility. The supplied scoliosis diagnosis and spinal procedure codes may also identify patients outside the intended cohort. The request specifies a 2006–2021 study period but supplies only ICD-10 (International Classification of Diseases) diagnosis codes. Codes like ICD and CPT (Current Procedural Terminology) can help find candidates, but may not represent the full intent; prior work has also found that ICD-based searches can miss musculoskeletal cases identified clinically [2].

**Figure 1:**
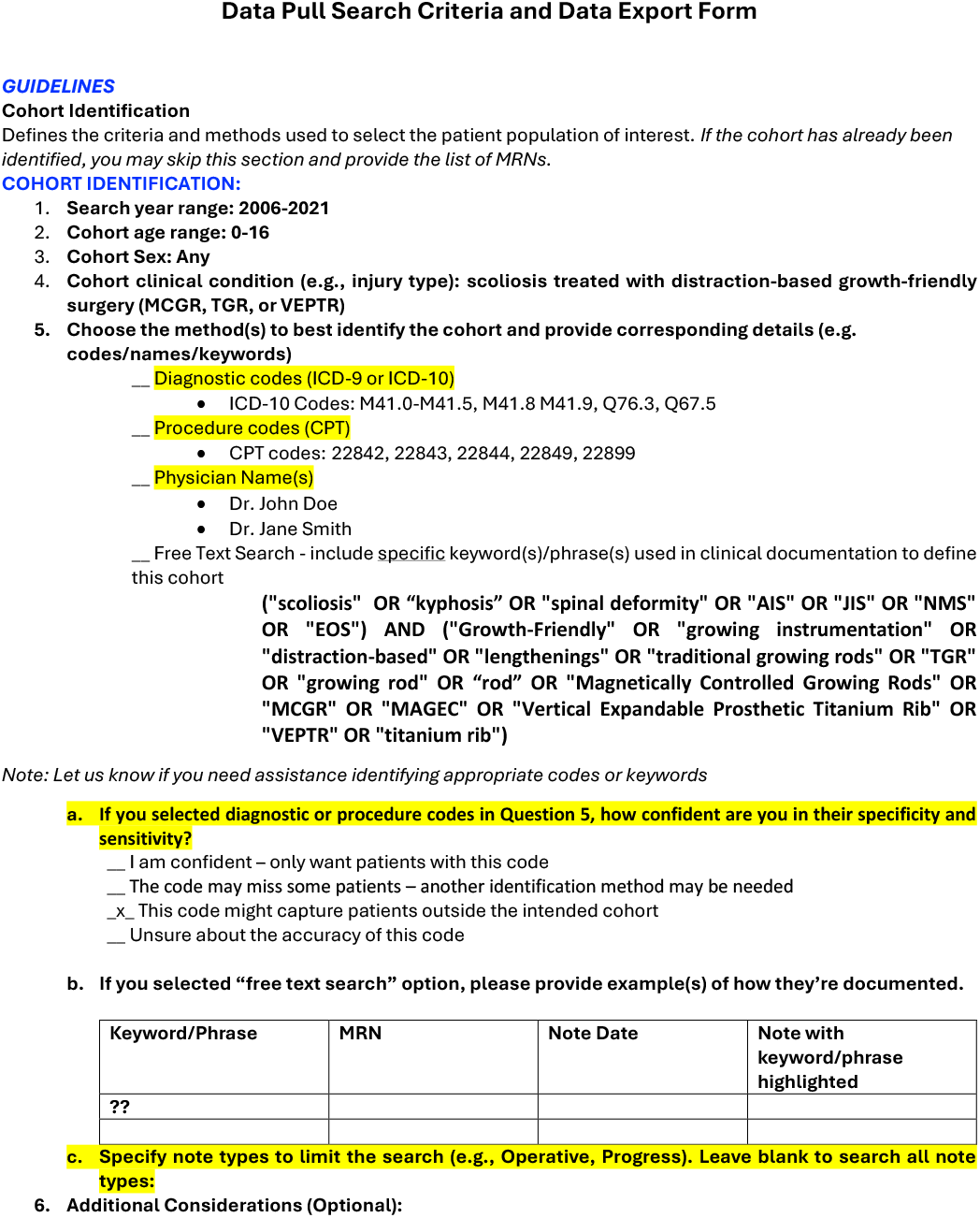
Representative data-query request illustrating incomplete correspondence between clinical intent and supplied structured search criteria. The defining treatment criterion—distraction-based growth-friendly surgery—is expressed in free text, while the requester indicates uncertainty in the specificity of the supplied ICD/CPT codes.

These requests are operational study specifications and not singular database queries. Codes may identify candidates, while final eligibility may depend on evidence from several EHR sources. Selfservice tools such as Epic SlicerDicer, i2b2, and Leaf[3] can reduce the need for custom programming, but users still need training and data literacy. Boolean logic, time relationships, and differences in how EHRs record clinical concepts can remain difficult to represent. Research staff, informaticians, and data analysts therefore continue to play an important role [4, 5].

Large language models (LLMs) have been used to generate EHR queries, plan multi-step database tasks, draft phenotype algorithms, and identify cohorts. TREQS and EHRSQL translate questions into queries over structured EHR tables; EHRAgent adds planning and query repair using execution feedback [6, 7, 8, 9]. Other studies generate phenotype algorithms or convert trial criteria into queries for standardized data models. Their evaluations report problems such as overly broad rules, restrictive rules, and incorrect concept mapping [10, 11]. A retrieval-augmented system demonstrated feasbility for cohort generation from observational study criteria reported an F1 score of 0.75, albeit on a smaller OMOP dataset with synthetically generated input requests with LLMs [12].

Prior LLM work also processes unstructured clinical notes. PRISM matches trial criteria to realworld oncology notes, and a recent Mass General Brigham preprint uses retrieval-augmented LLMs to identify dementia from EHR notes [13, 14]. These studies show that LLM-based note interpretation is not new, although they focus on a defined trial or phenotype. Only limited deployment in a production setting for such a solution has been reported: ClinQueryAgent was used in beta testing by staff across 15 NHS practices, and Ascent is described as deployed at Bayer for expertsupervised analysis of real-world clinical data [15, 16]. Prior work therefore spans benchmark datasets, institution-specific EHR evaluations, and early operational use; however it is limited to open datasets or offline experiments. Our study focuses on real investigator-submitted research requests that require routing across local codes, notes, and imaging, followed by request-specific free-text and unstructured note review. Table 1 summarizes these complementary approaches.

**Table 1:** Selected systems related to clinical research automation.

| **Approach** | **Method and evaluation** | **Distinctive scope and deployment evi-dence** |
|---|---|---|
| Text-to-SQL / agents [6, 7, 8, 9] | TREQS and EHRSQL translate questions into SQL over structured EHR tables; EHRAgent adds multi-step planning and query repair across three EHR datasets. | Evaluates question-to-query performance and tabular reasoning on benchmark databases; production deployment is not reported. |
| Phenotype / cri-teria SQL [10, 11] | Studies draft CDM phenotype queries or convert trial criteria to OMOP SQL. Evaluations include expert review, synthetic data, and selected vali-dation in an institutional OMOP database. | Translates predefined phenotypes or trial cri-teria into computable definitions; production deployment is not reported. |
| RAG cohort sys-tem [12] | Parses trial criteria, retrieves from specialized knowledge bases, maps concepts, and generates SQL; reports cohort-identification F1 of 0.75. | Closest criteria-to-cohort precedent, but evaluated on a defined research task; pro-duction deployment is not reported. |
| Note matching [13, 14] | PRISM matches trial criteria to real-world on-cology notes; a Mass General Brigham study uses RAG to identify dementia from EHR notes. | Addresses bounded trial or phenotype tasks rather than variable local-source routing. PRISM has reported clinical deployment; de-ployment of the dementia preprint is not re-ported [17]. |
| Operational agents [15, 16] | ClinQueryAgent reports beta use by 128 staff across 15 NHS practices; Ascent reports deploy-ment at Bayer for expert-supervised analysis of real-world clinical data. | Demonstrates operational use for population-health querying and real-world data analysis. |

The pipeline turns each natural-language data-query request into a set of executable EHR tasks. It routes request components to concept-specific agents and generates and corrects SQL using institution-specific templates prepared offline from historical queries, schema documentation, and database profiles. It then combines structured matches with relevant clinical notes and imaging evidence. For note review, a study-specific prompt guides the LLM to assess whether each note supports the request’s inclusion and exclusion criteria. The supporting evidence is retained for review, and note-level decisions are combined at the patient level. This lets the workflow assess clinical details that codes alone may miss, rather than treating a keyword match as proof of eligibility.

Our contributions include the following:

1. We present an agentic workflow that decomposes investigator requests into source-specific tasks, generates institution-specific SQL, and integrates structured EHR data with studyspecific clinical-note adjudication when required.
2. We present a expert-in-the-loop SQL template generation workflow to normalize institutionspecific EHR database schema for LLM context ingestion.
3. We evaluate the pipeline with various cohorts created with conventional methods like data analysts, self-service tools for discovery of patients in clinical research studies.

## Results

### Evaluation Requests

The evaluation included 50 pediatric orthopedic clinical data query requests submitted between mid-2024 and mid-2026 (Table 2). The requests covered a diverse range of orthopedic research studies including spine deformity, sports medicine, hip preservation, trauma, hand surgery, congenital disorders, guided growth, and other pediatric musculoskeletal conditions (Figure 2).

**Table 2:**
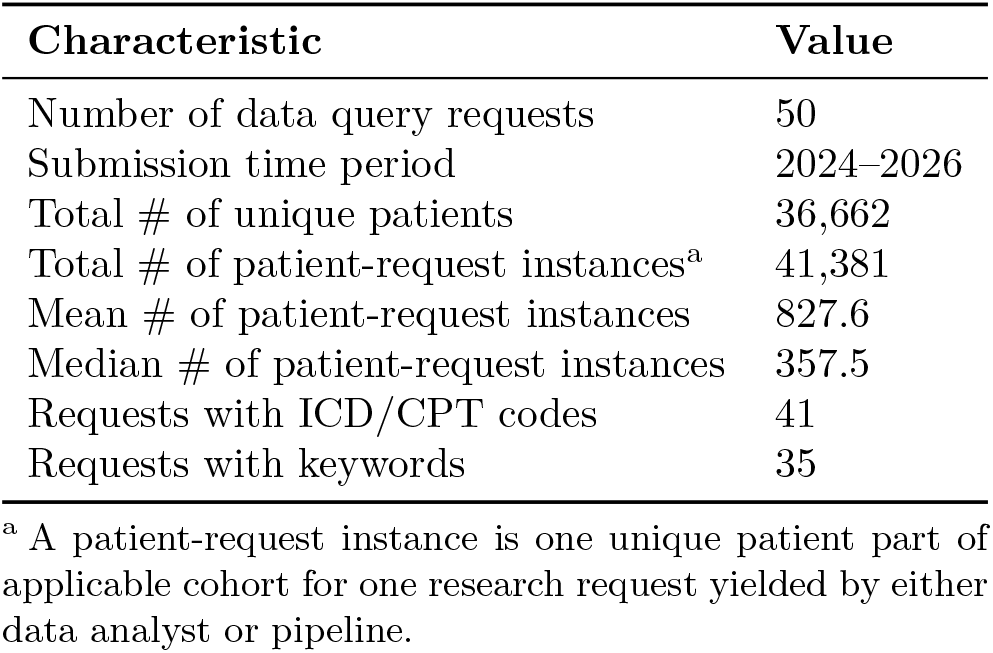
Study-Level Data Query Requests and Characteristics.

**Figure 2:**
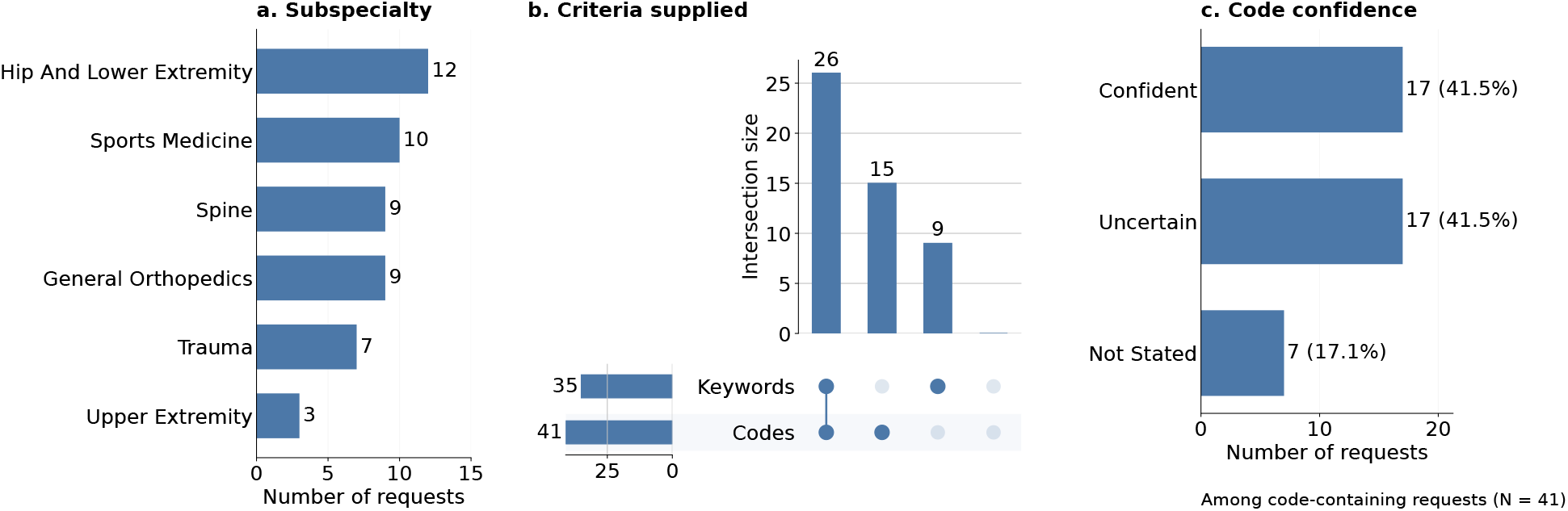
Characteristics of the evaluation set. (a) Distribution by orthopedic subspecialty. (b) UpSet plot of supplied search criteria: each vertical bar shows the number of requests in one exact combination, indicated by the dot matrix below (filled dots mark criteria present, connected dots mark criteria included together, and pale dots mark criteria absent) (c) Requester-reported confidence in supplied codes among the 41 code-containing requests

The pipeline supports three retrieval sources: billing codes (ICD/CPT), clinical notes, and radiology data. We selected these as an initial scope because they were frequently represented in the submitted requests. Diagnosis and/or procedure codes were supplied for 41 requests (82%), and 35 requests (70%) included note-search keywords. Imaging data is supported when a request requires radiological evidence. Extending the pipeline to support other domains, including medications, laboratory values, and pathology, is left to future work. Among code-containing requests with recorded confidence, half indicated that the supplied codes alone might not identify the intended population. These patterns required different combinations of structured retrieval, note searches, and clinical interpretation (Figure 2).

Manual review was completed for 43 requests. Seven were excluded from adjusted performance analyses: five were not selected for review because the workflows were highly concordant and/or the request specified only code-defined i.e., PPV and Sensitivity ≈ 0.99; one had incomplete eligibility criteria; and one required unsupported longitudinal reasoning. We report adjusted performance only for the 43 requests with completed review.

### Comparison with Data Analyst Cohorts

Historical cohorts created by expert data Analysts served as the comparator. For the 43 reviewed requests, annotators assessed random samples of patients identified by only one workflow. Estimated eligibility rates were applied to the corresponding discordant cohort in the adjusted performance calculations. The adjustment and its assumptions are described in Methods.

Manual review identified eligible patients in some pipeline-only cohorts and ineligible patients in some historical-only cohorts. After adjustment, median per-request Positive Predictive Value (PPV) was 0.93 (IQR, 0.82–0.99; 95% CI, 0.88–0.98), sensitivity within the observed union was 0.92 (IQR, 0.88–0.95; 95% CI, 0.90–0.95), and F1 score was 0.90 (IQR, 0.84–0.96; 95% CI, 0.87–0.94) (Figure 3a).

**Figure 3:**
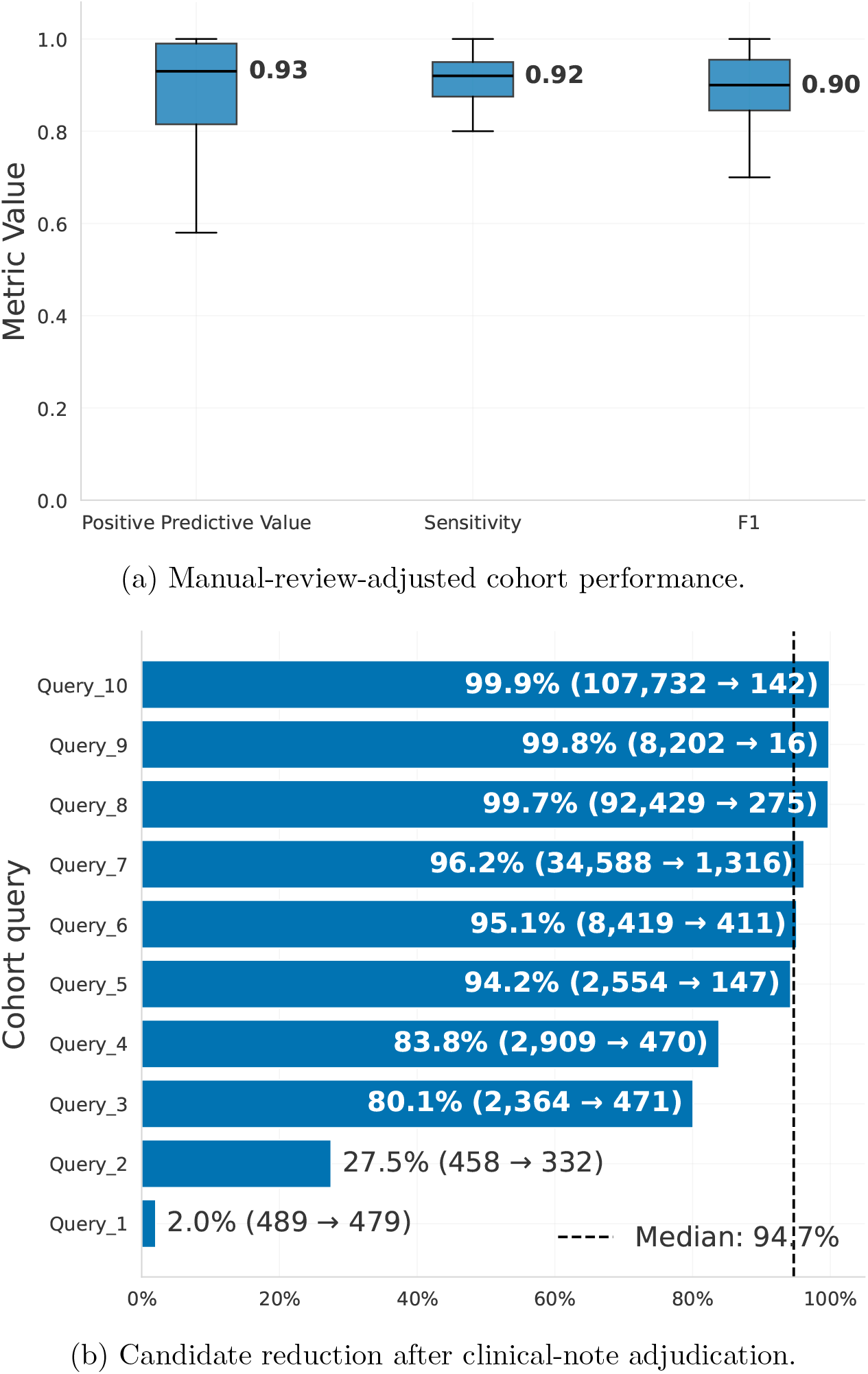
Cohort-identification performance and candidate reduction. (a) Positive predictive value, sensitivity within the observed union, and F1 score among 43 manually reviewed requests. Boxes represent interquartile ranges, horizontal bold lines indicate medians, and whiskers extend to the most extreme observations within 1.5 times the IQR. (b) Reduction in unique keyword-retrieved candidate patients for ten adjudicated requests without supplied ICD/CPT codes. Each bar represents one request; labels show percentage reduction and patient counts before and after adjudication. The dashed line indicates the median reduction (94.7%).

Across the annotated requests, the median per-request proportion of sampled pipeline-only patients judged eligible was 80.0% (IQR, 56.0%–95.0%). For sampled historical-only patients, the median per-request proportion judged ineligible was 33.3% (IQR, 18.7%–57.1%). These sample-based eligibility estimates were applied to the corresponding discordant patient cohorts when calculating adjusted performance metrics (Methods).

### Comparison with Conventional Methods

For this comparison, conventional methods were defined as candidate retrieval using diagnosis and/or procedure codes (ICD/CPT), where supplied, and keyword searches of clinical notes. This comparator was intended to approximate the initial candidate-identification stage of staff-led workflows using self-service tools such as Epic SlicerDicer or TriNetX.

For 10 of the 50 requests, selected randomly within the subset of queries where codes and/or keywords were provided, we performed additional manual review of patients retrieved by these conventional methods but subsequently rejected by the pipeline during note-level cohort adjudication. This review assessed whether the excluded patients were indeed ineligible or whether the adjudication stage had incorrectly excluded eligible cohort members, providing a check on the validity of narrowing the initial candidate sets.

Across these 10 requests, human annotators reviewed 588 patient–request instances excluded by the pipeline. The median per-request proportion of reviewed patients judged eligible by the annotators was 0% (IQR, 0.0%–1.5%). A near-zero proportion supports the validity of the pipeline’s note-level exclusion decisions among the reviewed patients.

We also quantified candidate reduction for ten adjudicated requests without supplied diagnosis or procedure codes. For each request, the candidate set comprised unique patients identified by clinical-note keyword retrieval. Reduction was calculated as 100×(*N*_candidate_ ™ *N*_retained_)*/N*_candidate_, where *N*_candidate_ is the number of unique patients retrieved before note-level adjudication and *N*_retained_ is the number retained afterward. The median per-request reduction was 94.7% (IQR, 81.0%–98.8%), corresponding to approximately 19 candidate patients for every patient retained (Figure 3b).

## Discussion

The main contribution of this study is demonstrating the feasibility of an agentic end-to-end pipeline to handle clinical research free-text requests made in a clinical research setting. The pipeline does more than generate SQL, it is capable of using LLM to ingest the free-text request, reason through selecting the required evidence sources, clinical concepts, generating SQL for querying the EHR, and perform adjudication on unstructured clinical notes specific to the inclusion/exclusion criteria. We validated this broader operationalization process through cohort identification, a common first step that requires the resulting workflow to produce a meaningful output.

The pipeline also addresses an important operational constraint in EHR-based clinical research. Eligibility screening requires substantial staff effort, and the burden increases when each candidate record must be reviewed manually [18]. A multi-stage workflow can change the allocation of that effort: structured retrieval can search broadly, request-specific note interpretation can resolve explicitly documented criteria, and clinical research staff can concentrate on ambiguous cases and final quality control. The reduction in keyword-retrieved candidates (Figure 3b) suggests a potential decrease in subsequent manual chart review.

Reviewing every patient in every cohort was not feasible. Instead, blinded review of sampled disagreements showed why historical cohorts should not be assumed to represent eligibility perfectly. Keeping the clinical evidence used for each decision makes disagreements easier to audit and supports focused human review. Prior phenotyping work has likewise shown the value of chart review for finding errors in both algorithms and reference cohorts [19].

The 50 requests spanned several pediatric orthopedic subspecialties and varied in codes, note terminology, imaging, demographics, and exclusions. This tested the workflow on distinct research requests rather than repeated runs of one phenotype. Reusable templates may preserve local EHR expertise and reduce the need to start each request from scratch. However, they also reflect local schemas and workflows. Prior work on phenotype portability shows why each new institution needs local validation [20].

Certain limitations constrain interpretation. This was a retrospective, single-institution evaluation in pediatric orthopedics. Historical Analyst-generated cohorts were imperfect comparators, and the adjusted estimates extrapolated validity rates from sampled disagreements to the full discordant sets. The estimates consequently depend on the size and representativeness of the annotated samples and do not establish absolute sensitivity in the underlying patient population. The current implementation is also limited to supported EHR concepts and retains a retrieval ceiling: incomplete requests, ambiguous terminology, unsupported longitudinal relationships, and clinical facts recorded outside the available EHR can affect the generated workflow and its outputs.

Future evaluation should test the workflow prospectively and across additional institutions, specialties, and classes of research request. Extensions to additional data domains, longitudinal reasoning, structured variable extraction, and uncertainty handling could broaden the range of requests that can be operationalized while directing human review toward ambiguous or conflicting cases. Taken together, the present findings support the feasibility of translating natural-language clinical research requests into executable, multi-stage automated workflows. Cohort identification provides an initial end-to-end validation of that capability; it does not establish that autonomous request operationalization can replace clinical oversight.

## Online Methods

### Input

#### Data Query Document

The present study was reviewed by the Boston Children’s Hospital Institutional Review Board, who determined that it met the criteria for exemption and waived the requirement of obtaining informed consent (IRB-P00046914). All methods were conducted in accordance with relevant guidelines and regulations. Following approval, we obtained requests submitted by clinical research staff (CRS) using a standardized Data Query Search Criteria Form (an example is shown in Figure 1). The form records the study period, age and sex restrictions, condition of interest, and requested retrieval methods. These may include ICD-9/10 diagnosis codes, CPT procedure codes, providers, and keywords or phrases expected in clinical documentation. The form also asks researchers to rate confidence in the codes, provide examples of note keywords, specify note types, estimate cohort size, and describe other inclusion or exclusion criteria.

#### Template Generation

An SQL template is a reusable, concept-specific query pattern that reflects the instiution’s EHR schema and querying conventions. Before evaluation, a data analyst uses the AI chat agent to prepare these templates through the three stages shown in Figure 4a.

**Figure 4:**
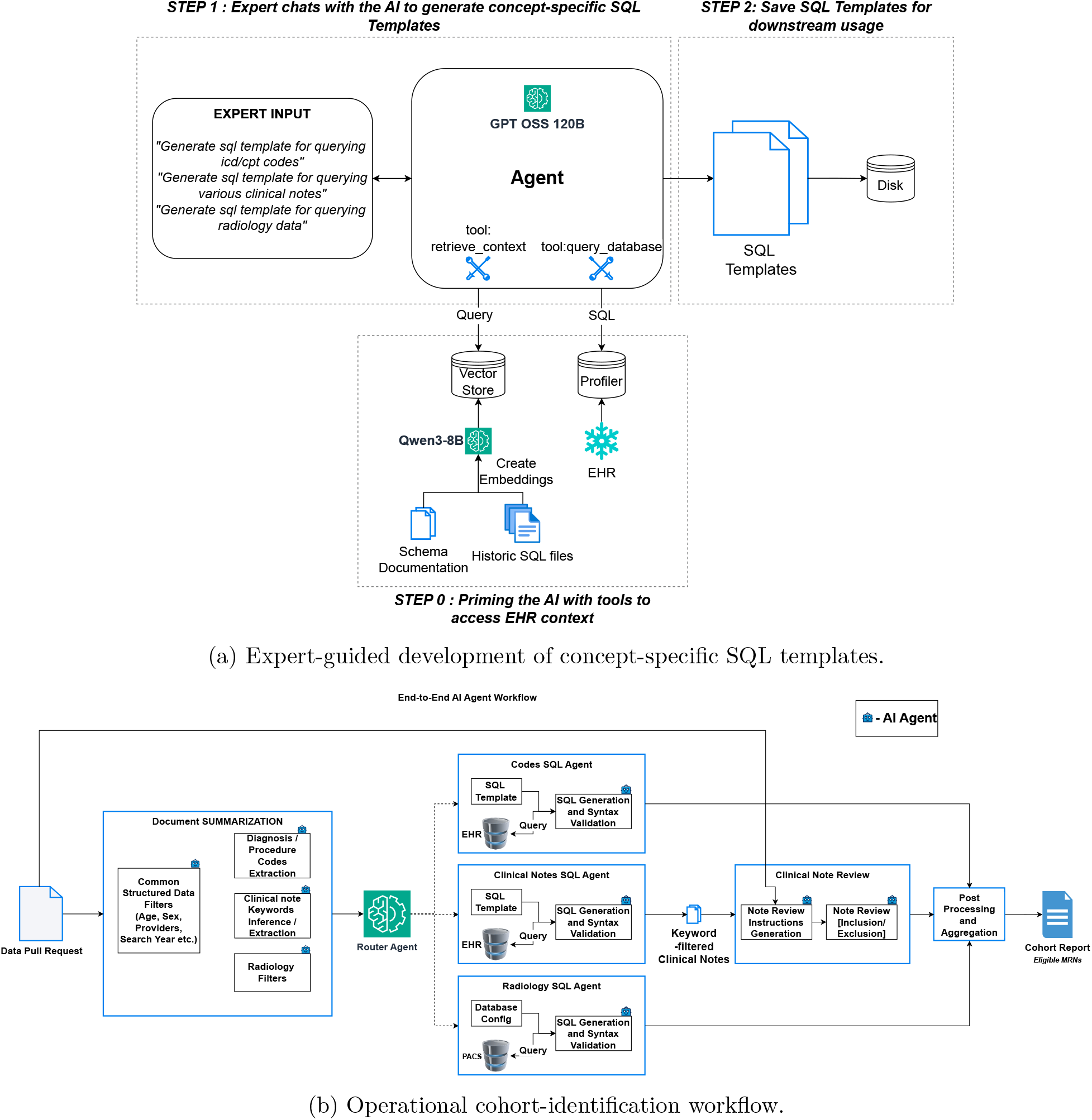
System development and execution. Panel (a) shows the three-stage preparation of SQL templates: context assembly, template drafting with expert feedback, and saving templates for downstream use. Panel (b) shows how investigator requests are summarized and routed to specialized retrieval agents, followed by note-level review, patient-level aggregation, and cohort reporting.

##### Step 0: Prepare local EHR context

The figure’s retrieve_context and query_database tools provide the AI agent to selectively choose the specific information it needs to answer the user’s request for a specific SQL template. The development sources for these tools include:

1. A repository of over 100 historical SQL queries created by three data analysts between 2025 and 2026
2. Internal EHR documentation maintained by the clinical informatics team describing data concepts and standard querying practices
3. Table-profile information containing column-level details such as

- Data type
- Data sparsity
- Top-*k* values (*k* = 7)
- Random-*k* values (*k* = 7)

The historical SQL queries and internal EHR documentation were divided into chunks, embedded using Qwen3-8B[21], and stored in a vector store.

##### Step 1: Draft concept-specific templates with expert feedback

The interaction along with verification from the expert user yields a concept-specific template informed and grounded in relevant examples and documentation from the EHR context store. The expert can provide feedback during this interaction to refine the template for the local schema and querying practices. GPT-OSS-120B[22] assisted in drafting and refining the templates using this retrieved context.

##### Step 2: Save templates for downstream use

The resulting templates were saved to disk and fixed before evaluation. During request processing, the SQL-generation agent used the corresponding stored template as context to generate a request-specific query. Supplementary Listings S1–S3 show representative queries for code retrieval, note retrieval, and encounter-level variables

### SQL Generation Pipeline

Figure 4b shows the workflow. Specialized agents route the request, summarize its criteria, generate and correct SQL, create a note-review prompt, and integrate the results. Supplementary Figure S1 shows the corresponding routing graph.

The router reads the data-query request and selects the relevant retrieval domains. Its system instructions are provided in Supplementary Listing S4.

For each selected domain, a summary agent extracts the information needed for retrieval. The code agent records ICD/CPT codes and related criteria. The note agent identifies diagnosis or procedure descriptions and suggests terms likely to appear in notes. The radiology agent extracts details such as modality, body region, and imaging features. Each summary also includes applicable age, sex, and study-period filters. Instructions for these agents are in Supplementary Listings S6–S8.

The summarized retrieval objective is then passed to a concept-specific SQL generation agent, together with the corresponding SQL template from the template repository described under Template Generation. Each generated SQL query underwent an automated compilation check using the Snowflake EXPLAIN command before execution. Queries that produced syntax or compilation errors were returned to the SQL-generation agent together with the database error message, and the agent iteratively revised the query until compilation succeeded or the retry limit(*n* = 3) was reached. This step verifies that the generated SQL is syntactically valid and can be compiled by the database engine. The code, clinical-note, and radiology SQL-generation instructions are provided in Supplementary Listings S9–S11, and the SQL validation instructions are provided in Supplementary Listing S5.

### Note-level Cohort Adjudication

Some eligibility criteria are documented in clinical notes rather than structured fields. Examples include diagnostic confirmation, syndromic variants, operative findings, device use, laterality, and recurrence. Codes alone may not capture these details.

For requests that require note review, a prompt-generation agent converts the request’s inclusion, exclusion, and extraction criteria into a study-specific prompt.

The prompt is applied to the retrieved notes to assess whether their documented evidence meets the request criteria. Prompt-generation and note-review instructions are provided in Supplementary Listing S12.

### Cohort construction and branch integration

The request form asks whether billing codes alone define the cohort or whether clinical-note review is also required. This choice determines where the search begins. For a codes-only request, matching diagnosis and/or procedure records generate the candidate cohort; note-level LLM review is not performed. For a request that requires note review, keyword-filtered notes generate candidates. If codes are also supplied, code matches are added as supporting evidence, not used as a required gate. If no codes are supplied, the note search and adjudication still proceeds.

When radiology is requested, imaging results are linked to relevant notes by date. The default window is within one year before or after the note date. The study-specific prompt evaluates each retrieved note against the request’s inclusion and exclusion criteria. A patient is retained if at least one note meets all required criteria. Code matches and linked imaging evidence are considered during review, and the final cohort is deduplicated by patient.

Age, sex, provider, and study-period filters are applied to the relevant search branches. Requests that require unsupported data or reasoning—such as medication, laboratory, or pathology retrieval, or complex longitudinal sequencing—are flagged as out of scope by the router agent.

### Implementation

The pipeline used LangGraph, with LangChain for orchestration. Agents handled routing, request summarization, SQL generation and correction, prompt generation, note review, and result integration. During evaluation, all agents used GPT-OSS-120B; Qwen3-8B was used for historical-query retrieval during offline template development (see Template Generation). No task-specific finetuning was performed. Model inference used vLLM on hardware comprising eight NVIDIA H200 GPUs, with temperature 0 and top-*p* 1.0 for LLM calls.

Microsoft Word (.docx) request forms were converted to Markdown with Docling. SQL queries that passed compilation checks were executed against the institutional Snowflake EHR using Python’s Snowflake connector.

### Blinded manual review of discordant patients

Two health informatics students performed the manual review. Requests were divided between them, so each request was reviewed by one annotator. For each sampled patient, the annotator received the original request and the clinical evidence used by the workflow. The request served as the eligibility rubric. Annotators applied each study’s diagnosis, procedure, demographic, time, inclusion, and exclusion criteria; no single rubric was used across all requests.

Annotators were blinded to whether a patient came from the pipeline-only or historical-only group. They assigned a binary label indicating whether the evidence supported inclusion. Because patients were not independently reviewed by both annotators, inter-rater agreement was not measured.

### Reference cohort and performance metrics

Manual review was completed for 43 requests. For each request *r*, let *P* denote the set of patients identified by the proposed pipeline and *H* the historical cohort generated by the expert data analyst workflow. Patients were partitioned into three observable patient cohorts: pipeline-only patients *A* = *P* \ *H*, historical-only patients *B* = *H* \ *P*, and patients identified by both workflows *C* = *P* ∩ *H*. Patients identified by neither workflow, *D* = Ω \ (*P* ∪ *H*), were not observable within the comparison design. For *r* and discordant cohort *g* ∈ {*A, B*}, let *N*_*r,g*_ be the number of patients in that group. A random sample without replacement from *N*_*r,g*_ was selected using these sample sizes:

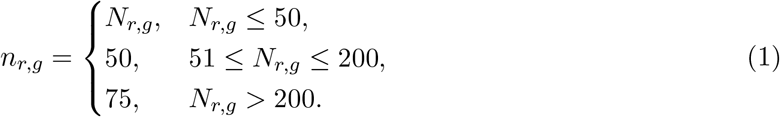

The thresholds and conditional sample size values was decided taking into account the availability and number of annotators and median size of the discordant patient cohorts.

The blinded sample review estimated the proportions eligible in *A* and *B*, denoted by 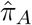 and 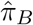.

Assuming patients in *C* were eligible, positive predictive value (PPV) was calculated as

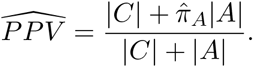

Sensitivity within the observed union of both cohorts was calculated as

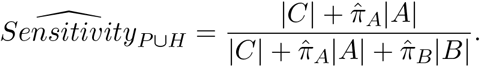

F1 was calculated from:

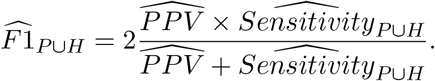

These measures describe pipeline performance among patients identified by at least one workflow. They do not estimate absolute sensitivity in the EHR population because patients missed by both workflows (*D*) were not observed. The estimates also depend on the assumption that all patients in *C* were eligible and on whether the sampled disagreements represent their full patient cohorts.

## Data Availability

The data used in this study cannot be publicly shared due to patient privacy concerns. Data can be accessed upon reasonable request pending approvals from Boston Children’s Hospital.

## Code Availability

Code to replicate the results of this paper can be accessed upon reasonable request pending approvals from Boston Children’s Hospital.

## Acknowledgments

We would like to acknowledge funding support from the Children’s Orthopedic Surgery Foundation and Hardware support from NVIDIA.

## Author Contributions

Concept and design: K.P., N.T., A.M.K. Acquisition, analysis, or interpretation of data: K.P., N.T., N.K. Drafting of the manuscript: K.P., N.T., A.M.K. Critical revision of the manuscript for important intellectual content: All authors. Statistical analysis: K.P., N.T., A.M.K. Obtained funding: A.M.K. Administrative, technical, or material support: N.T., A.M.K. Supervision: A.M.K., N.T. Manual annotation: A.K., M.B. Initial concept proposal and literature review: A.W.

## Competing Interests

The authors declare no competing interests.

## Notes

### Competing Interest Statement

The authors have declared no competing interest.

### Author Declarations

Institutional Review Board of Boston Children's Hospital gave ethical approval for this work (IRB#: IRB-P00046914).

### Summary of Updates

Fixed author name in references

